# Changing patterns of cigarette and ENDS transitions in the US: a multistate transition analysis of youth and adults in the PATH Study in 2015–17 vs 2017–2019

**DOI:** 10.1101/2022.12.15.22283292

**Authors:** Andrew F. Brouwer, Jihyoun Jeon, Evelyn Jimenez-Mendoza, Stephanie R. Land, Theodore R. Holford, Abigail S. Friedman, Jamie Tam, Ritesh Mistry, David T. Levy, Rafael Meza

## Abstract

**Introduction:** It is unknown how recent changes in the tobacco product marketplace have impacted transitions in cigarette and electronic nicotine delivery system (ENDS) use.

**Methods:** A multistate transition model was applied to 24,242 adults and 12,067 youth in Waves 2–4 (2015–2017) and 28,061 adults and 12,538 youth in Waves 4–5 (2017–2019) of the Population Assessment of Tobacco and Health (PATH) Study. Hazards for initiation, cessation, and product transitions were estimated in multivariable models, accounting for gender, age group, race/ethnicity, and daily vs non-daily product use.

**Results:** Changes in ENDS initiation/relapse rates from never, non-current, and cigarette-only use depended on age group, including among adults. Among youth never users, the 1-year probability of ENDS initiation increased after 2017 from 1.6% (95%CI: 1.4-1.8%) to 3.8% (95%CI: 3.4-4.2). Persistence of ENDS-only use (1-year probability of remaining an ENDS-only user) increased for youth from 40.7% (95%CI: 34.4-46.9%) to 65.7% (95%CI: 60.5-71.1%) and for adults from 57.8% (95%CI: 54.4-61.3%) to 78.2% (95%CI: 76.080.4%). Persistence of dual use similarly increased for youth from 48.3% (95%CI: 37.4-59.2%) to 60.9% (95%CI: 43.0-78.8%) and for adults from 40.1% (95%CI: 37.0-43.2%) to 63.8% (95%CI: 59.6-67.6%). Youth and young adult dual users became more likely to transition to ENDS-only use but middle-aged and older adults did not.

**Conclusions:** ENDS and dual use have become more persistent. Middle-aged and older adult dual users have become less likely to transition to cigarette-only use but not more likely to discontinue cigarettes. Youth and young adults are more likely to transition to ENDS-only use.

## Introduction

Electronic nicotine delivery systems (ENDS), including e-cigarettes, have substantially changed the landscape of tobacco and nicotine products in the US and other countries. In the US, ENDS sales, driven by JUUL and similar products, increased dramatically over 2018 [1–3] and fueled concerns about a potential youth vaping epidemic [4]. ENDS, as an alternative to traditional cigarettes, may create a public health benefit through harm reduction if cigarette users leverage ENDS to quit or reduce smoking or if ENDS divert would-be cigarette users into ENDS use [5–8]. However, the real-world impact of ENDS is uncertain, as it is unclear whether ENDS facilitate smoking cessation broadly or whether they inhibit long-term smoking cessation through continued nicotine addiction [9–11]. Moreover, concerns about ENDS use by youth leading to smoking initiation remain, despite continued declines in youth smoking [12]. The impact of ENDS may depend on the product generation. Unlike newer ENDS products using nicotine salts, early freebase nicotine ENDS products were unpalatable at higher nicotine concentrations, so that different products may have substantially different impacts on tobacco use behaviors [13]. These newer products also debuted new flavors, many of which appeal to youth [14]. Accordingly, it is important to investigate how transitions between tobacco products (initiation, cessation, product switching) changed after 2017 and how these changes depend on age group. This analysis focuses on cigarettes—the most used combustible tobacco product—and ENDS—which capture a variety of nicotine vaping products—to better understand the potential public health impact of changes in the marketplace on smoking and vaping.

Interdependencies make it difficult to interpret changes in the fraction of people transitioning from one type of product to another, or to non-use. For example, an increase in the *rate* that cigarette-only users transition to dual use of cigarettes and ENDS inherently decreases the *fraction* of cigarette-only users that quit, even if the quitting rate is unchanged, because there are fewer cigarette-only users left to quit. Accordingly, we need to estimate the transition rates that underly the observed transition fractions to be able to attribute changes in the fractions to changes in propensities to use or quit products. To do so, we use multistate transition models, which are increasingly being used to analyze tobacco product transitions and to understand how sociodemographic factors impact these underlying rates [15–19]. In a previous multistate transition analysis, we found that ENDS use in 2013–2017 was less persistent than cigarette use in the US [15], but more recent work has suggested that ENDS use has become more persistent in recent years [20]. In this analysis, we implemented a multistate transition model using data from the Population Assessment of Tobacco and Health (PATH) Study, distinguishing between Waves 2–4 (2015–2017) and Waves 4–5 (2017–2019) for adults and youth, and adjusting for demographics and frequency of product use. We also developed an approach to calculate continuous, spline-based estimates of the impact of age on tobacco product transitions, allowing a clearer picture of exactly how the rate of each transition varies by age and how these patterns changed after 2017.

## Methods

### Data

The Population Assessment of Tobacco and Health (PATH) Study is a national longitudinal cohort study of tobacco and nicotine product use behaviors among the civilian noninstitutionalized adult (ages 18–90) and youth (ages 12–17) populations [21]. The initial, nationally representative Wave 1 cohort was replenished in Wave 4 to form the nationally representative Wave 4 cohort (about 25% of the Wave 4 cohort were not in the Wave 1 cohort). Our analysis compared 24,242 adults and 12,067 youth from the Wave 1 cohort in Waves 2–4 (Oct 2014 to Jan 2018, abbreviated as 2015–2017) and 28,061 adults and 12,538 youth in the Wave 4 cohort in Waves 4–5, including Wave 4.5 for youth (Dec 2016 to Nov 2019, abbreviated as 2017–2019). Shadow youth (i.e., youth under age 12 who were preemptively enrolled but did not participate in data collection) aged into the youth cohort in each Wave, maintaining the age distribution of youth that we follow to the subsequent wave. Transitions of youth who were age 17 were observed by determining their product use at age 18 in the adult cohort. Time between follow-up for each participant was approximately one year, except for the two-year gap for adults between Waves 4 and 5, as adults were not included in Wave 4.5. The Markov framework explicitly accounts for the time between observations, allowing for joint analysis of data with varying follow-up times. We used information on age (both as age in years and age groups 12–14, 15–17, 18–24, 25–34, 35–54, 55–90), gender (male, female), and race/ethnicity (Hispanic, non-Hispanic Black, non-Hispanic White, other/unknown).

Participants were classified as a never user, non-current user, cigarette-only user, ENDS-only user, or dual user, as in previous work [15]. In brief, never users reported no established use of cigarettes (<100 lifetime cigarettes) or ENDS (never “fairly regular” use), and the other states were defined based on both established use and past 30-day use of either or both products. Non-current use is defined as ever established use of either cigarettes or ENDS but no past-30-day use of either product. A summary of the state definitions is given in the supplementary material (Figure S1A). Additionally, participants using cigarettes or ENDS were classified as daily or non-daily users depending on whether they reported using the product 30 out of the past 30 days or fewer than 30 (based on previous analysis of the distribution of reported use [22]). Characteristics of the populations are given in the supplementary material (Table S1). This analysis was deemed exempt from regulation as human subjects research (University of Michigan Institutional Review Board HUM00162265).

### Transition modeling

We used a previously developed multistate transition model [15, 16, 23] to analyze the underlying transition hazard rates between product use and covariate hazard ratios for four different groups: youth 2015–2017 (Waves 2–4), youth 2017–2019 (Waves 4–5), adults 2015–2017 (Waves 2–4), and adults 2017–2019 (Waves 4–5). Multistate transition models are continuous time, finite-state stochastic process models that assume that transition hazard rates depend only on the current state and not on past states or transition history [24]. We incorporated Wave 1 and Wave 4 longitudinal survey weights into the model, as described in Brouwer et al. [15], which provides further technical details, and the included modeled transitions are given in the supplementary material (Figure S1B). The model estimates instantaneous risk of transition from one state to another, i.e., transition hazard rates, which collectively define the probability of transitioning from one state to any other at a future time. We separately estimate the transition rates for youth and adults in 2015–2017 (Waves 2–4) and 2017–2019 (Waves 4–5), adjusting for gender, age group, race/ethnicity, and daily vs non-daily cigarette and ENDS use, the effects of which are estimated as covariate hazard ratios (HR). We estimated the effect of a continuous age on the transition using cubic B-splines (a kind of piecewise polynomial) [25] with knots at ages 12, 18, 40, and 90.

## Results

Throughout the results, we compare transition probabilities, hazard rates, and hazard ratios between 2015–2017 (Waves 2–4) and 2017–2019 (Waves 4–5).

### Transition probabilities and hazard rates

Among youth never users, the 1-year probability of ENDS initiation increased from 1.6% (95%CI: 1.4– 1.8%) to 3.8% (95%CI: 3.4–4.2) (Figure 1A and B). Persistence of youth ENDS-only use increased from 40.7% (95%CI: 34.4–46.9%) to 65.8% (95%CI: 60.5–71.1%). Youth cigarette-only users became more likely to transition to dual use, with the 1-year transition probability increasing from 15.0% (95%CI: 8.6– 21.5%) to 23.9% (95%CI: 16.0–31.9). Concurrently, dual users became more likely to transition to ENDS-only use, with the transition probability increasing from 5.2% (95%CI: 0.8–9.6%) to 16.5% (95%CI: 6.7–26.3%).

**Figure 1:**
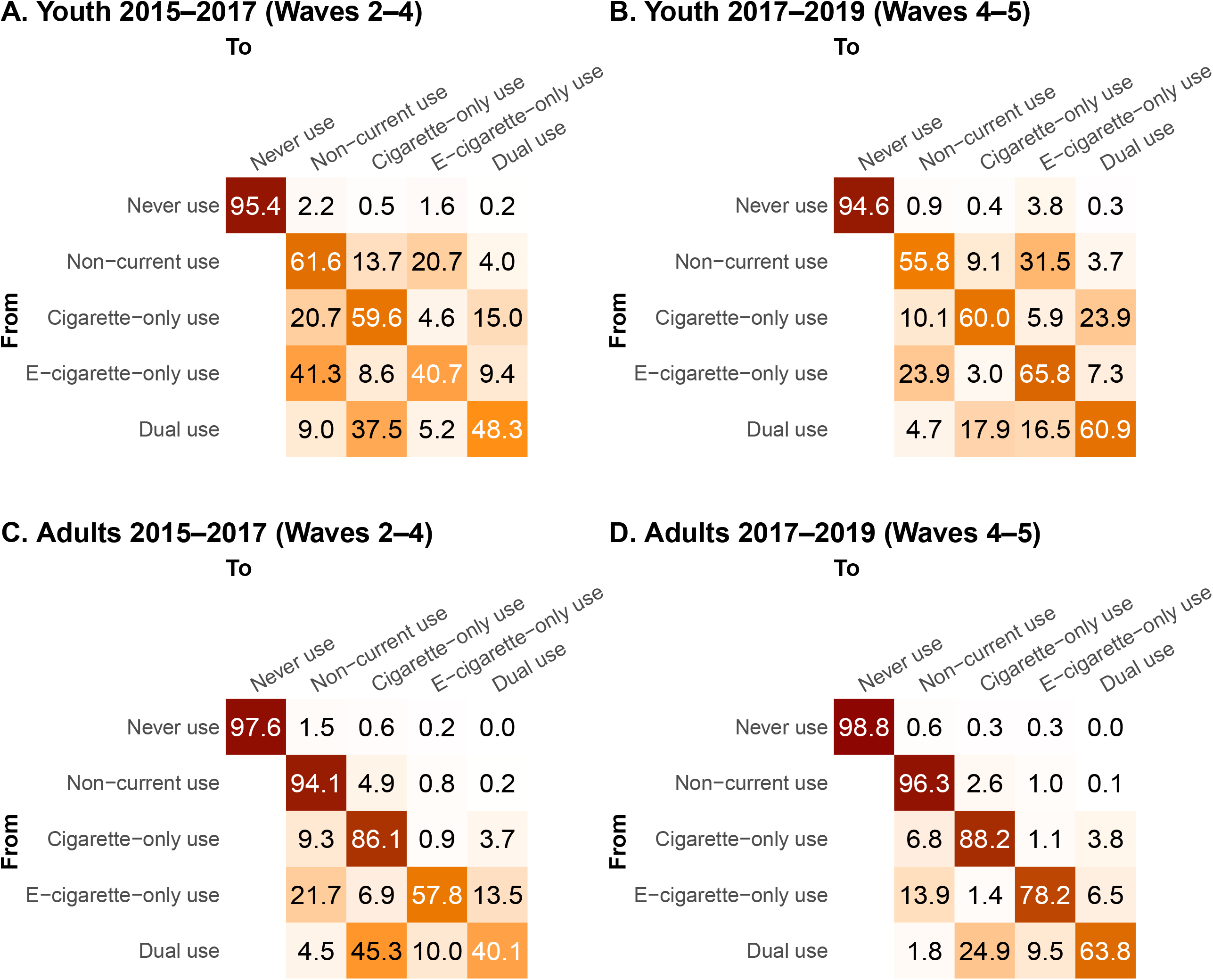
One-year transition probabilities for adults and youth in 2015–2017 (Waves 2–4) and 2017– 2019 (Waves 4–5). Confidence intervals are given in Figure S1.

For adults overall, there was little change after 2017 in transitions for cigarette-only users. Dual use became more persistent, with the 1-year probability of remaining a dual user increasing from 40.1% (95%CI: 37.0–43.2%) to 63.8% (95%CI: 59.9–67.6%)), accompanied by a decrease in transitions to cigarette-only use, which decreased from 45.3% (95%CI: 42.4-48.3%) to 24.9% (95%CI: 21.5,28.3%) (Figure 1C and D). One-year persistence of ENDS-only use also increased from 57.8% (95%CI: 54.4– 61.3%) to 78.2% (95%CI: 76.0–80.4%). For adult never users overall, there was no significant change in ENDS initiation, which remained low at 0.2% (95%CI: 0.2–0.3%) in 2015–17 and 0.3% (95%CI: 0.3– 0.4%) in 2017–2019.

Because the transition *probabilities*, which must sum to one, are interdependent, they do not directly indicate whether propensities to initiate, quit, or change products are increasing or decreasing. Instead, the underlying transition *hazard rates* reflect these propensities. We find that transition hazard rates decreased significantly among adults from 2015–2017 to 2017–2019, except for never or non-current to ENDS-only use (Figure 2A). That is, increases in ENDS and dual use persistence are driven by decreases in other transitions in the cohort while never or non-current to ENDS-only use transitions stayed constant. Among youth, fewer changes in the transition rates were statistically significant (Figure 2B). Exceptions include the increase in initiation from never to ENDS-only use and the decreases in the transition hazard rates from ENDS-only use to either non-current or dual use.

**Figure 2:**
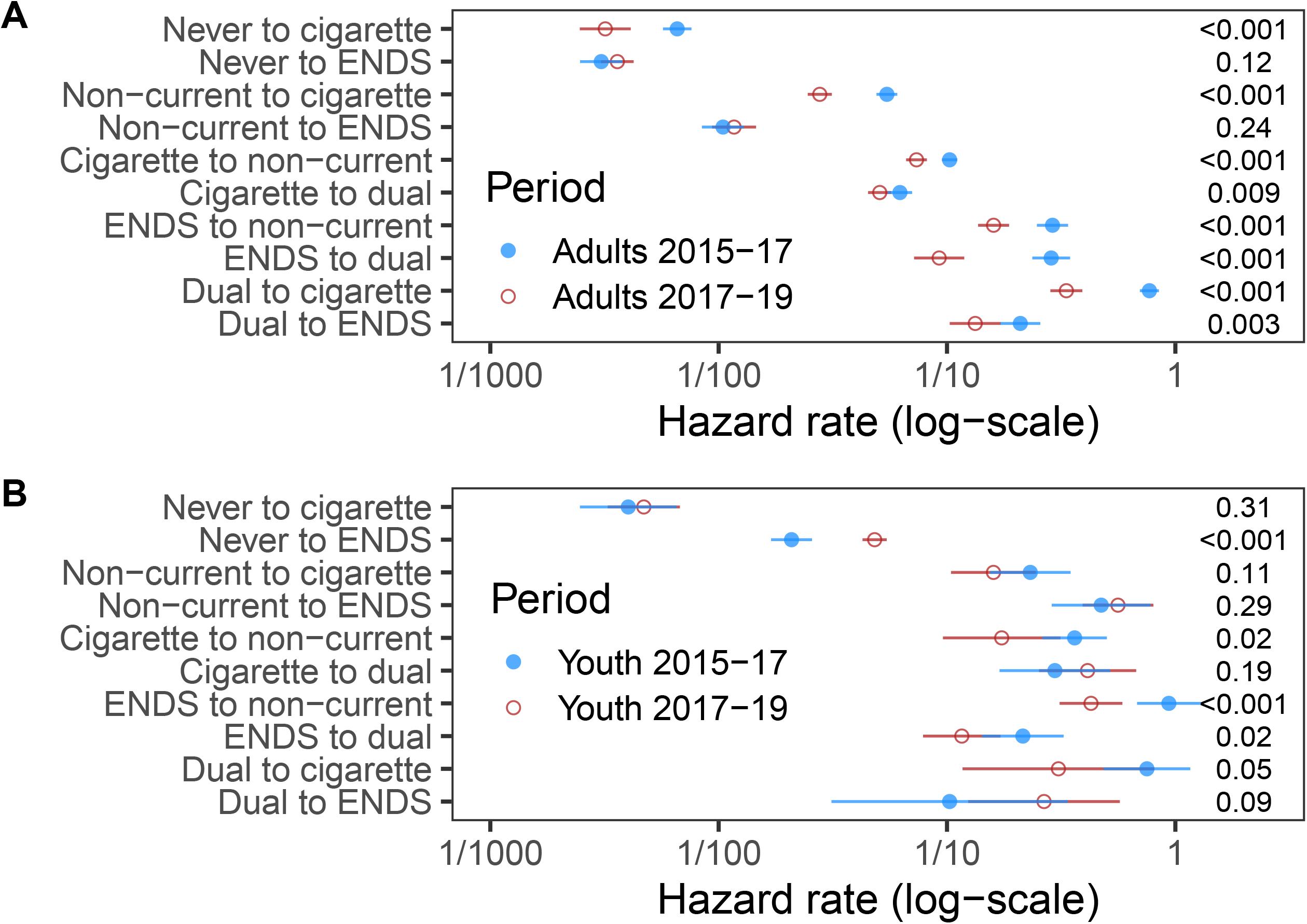
Transition hazard rates among adults and youth in 2015–2017 (Waves 2–4) and 2017–2019 (Waves 4–5). Values on the right-hand side are the p-values for difference in rates between the two periods.

### Sociodemographic transition hazard ratios

There were few changes in transitions patterns by gender or race/ethnicity for adults or youth, and the impacts of non-daily vs daily use of cigarettes or ENDS were often difficult to interpret because of high variance of the estimates in many instances (Table 2). Compared to youth ages 15–17, youth ages 12–14 had a lower initiation rate of cigarettes and ENDS from never use as well as a higher rate of ENDS cessation. However, the HRs comparing the two age groups (hazard ratios) for most transitions for youth did not change significantly from 2015–2017 to 2017–2019 even when the absolute transition rates did (Figure 2). Thus, differences in transition probabilities over time for the two youth age groups (Fig 3A-D) reflect changing absolute transition rates overall but stable relative transition rates.

**Figure 3:**
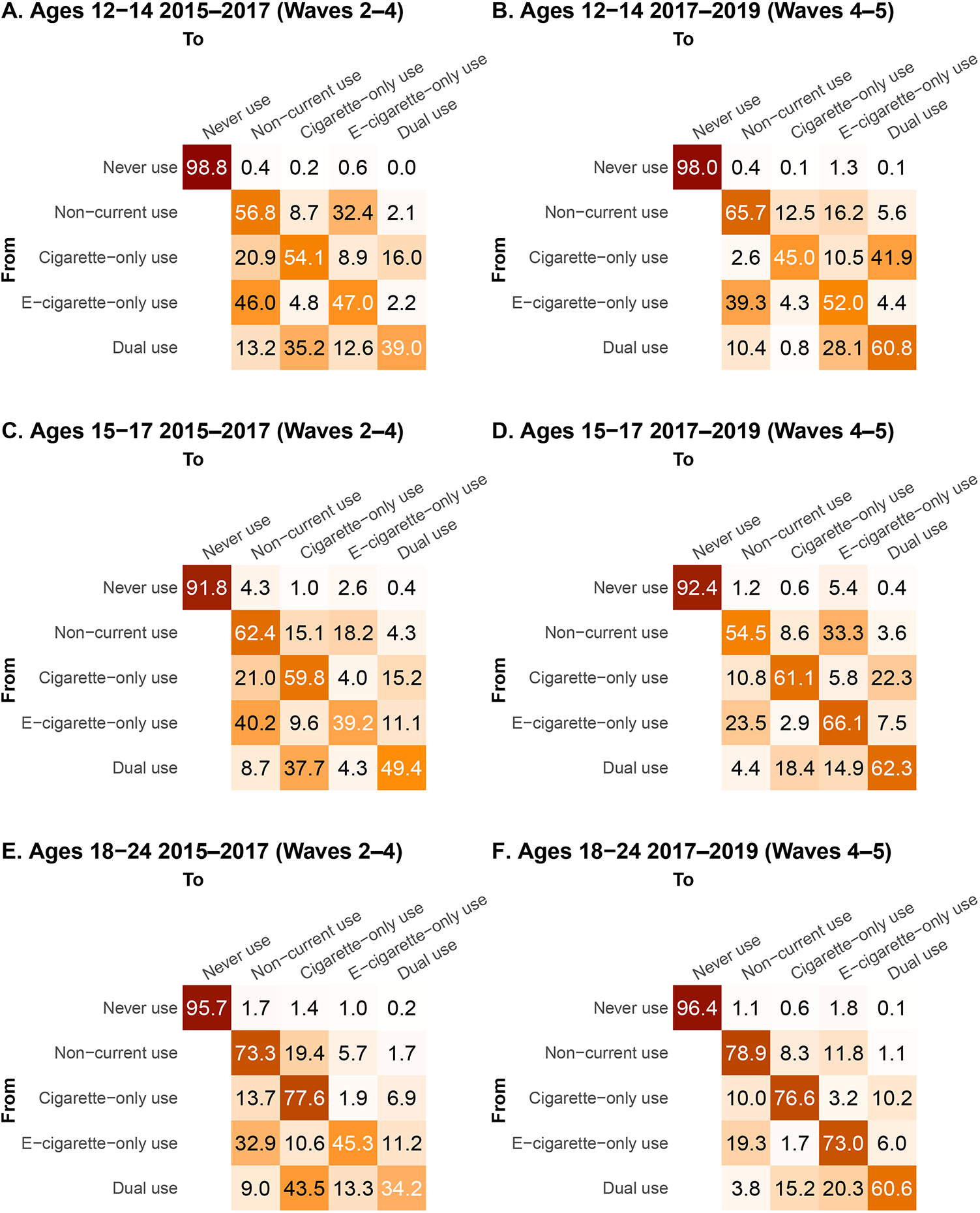
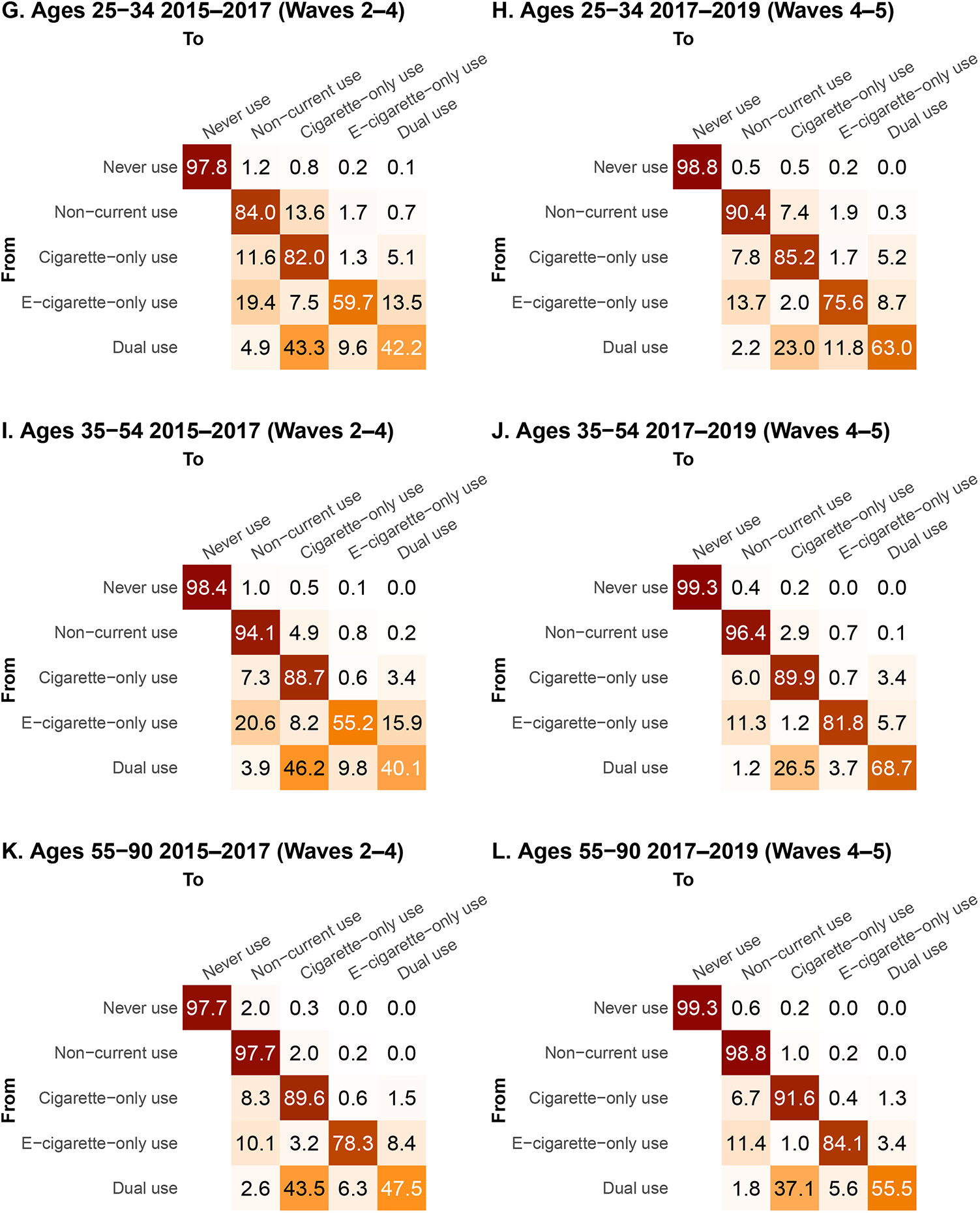
One-year transition probabilities by age group in 2015–2017 (Waves 2–4) and 2017–2019 (Waves 4–5). Confidence intervals are given in Figure S2.

Unlike for youth, HRs comparing adult age groups did change for several transitions. For example, there was a statistically significant increase in the HR of transitioning from non-current to ENDS-only use, with a HR of 9.48 (95%CI: 5.48–16.4) for ages 18–24 vs 35–54 in 2015–2017 compared with 22.2 (95%CI: 14.0–35.2) in 2017–2019 (p=0.01 for test of difference of means of the log hazard ratios). The change in the HR for transitions from never use to ENDS-only use for young adults (ages 18–24) vs middle-aged adults (35–54) had a large point estimate but was not statistically significant, with a HR of 20.5 (95%CI: 4.59–87.6) for ages 18–24 vs 35–54 in 2015–2017 compared with 82.2 (95%CI: 13.3–509) in 2017–2019 (p=0.12). Other hazard ratios are given in Table 2. The changing HRs by age drove the statistically significant increase in the probability of transitioning from non-current use to ENDS-only use among ages 18–24, accompanied by a decrease in the relapse to cigarettes (Figure 3E-F). Specifically, for ages 18–24, the probabilities of transitioning from non-current use to cigarette-only and to ENDS-only use, respectively, were 19.4% (95% CI: 16.5–22.3%) and 5.7% (95%CI: 4.1–7.3%) in 2015–2017 and 8.3% (95%CI: 6.7–9.9%) and 11.8% (95%CI: 9.4–14.2%) in 2017–2019. For ages 35–54 these transition probabilities were 4.9 (95% CI 3.9–5.8%) and 0.8% (95%CI: 0.4–1.1%) in 2015–2017 and 2.9% (95%CI: 0.7–1.3%) and 0.7% (95%CI: 0.4–0.9%) in 2017–2019.

The HR for ages 18–24 vs ages 35–54 for the transition from dual use to ENDS-only use also increased (p=0.005) from HR 1.32 (95%CI: 0.64–2.71) to 5.48 (95%CI: 2.40–12.5). These changes in the transition HRs resulted in differential changes in transition probabilities by age (Figure 3). Young adult (ages 18-24) dual users became much more likely to remain a dual user (34.2% to 60.6%), less likely to transition to cigarette-only use (43.5% to 15.2%), and marginally more likely to transition to ENDS-only use (13.3% to 20.3%), and much more likely to remain a dual user (34.2% to 60.6%). Middle-aged adults (ages 35-54) dual users also became more likely to remain dual users (40.1% to 68.7%) and less likely to transition to cigarette-only use (46.2% to 26.5%), but they became *less* likely to transition to ENDS-only use (9.8% to 3.7%).

### Impact of age on transition rates

Spline estimates of transition rates as a function of age in years rather than age group are given in Figure 4. ENDS initiation or relapse from never use, non-current use, or cigarette-only use (to dual use), occurs primarily among youth and young adults, peaking in the late teen years. The rate of transitioning from ENDS-only to non-current use is also higher among youth and young adults, compared to older individuals. Other transitions show comparatively smaller age differences. The transitions that changed the most over all ages are the ENDS-only to non-current use, ENDS-only to dual use, and dual to cigarette-only use transitions; rates for each of these transitions decreased for most ages (Figure 4).

**Figure 4:**
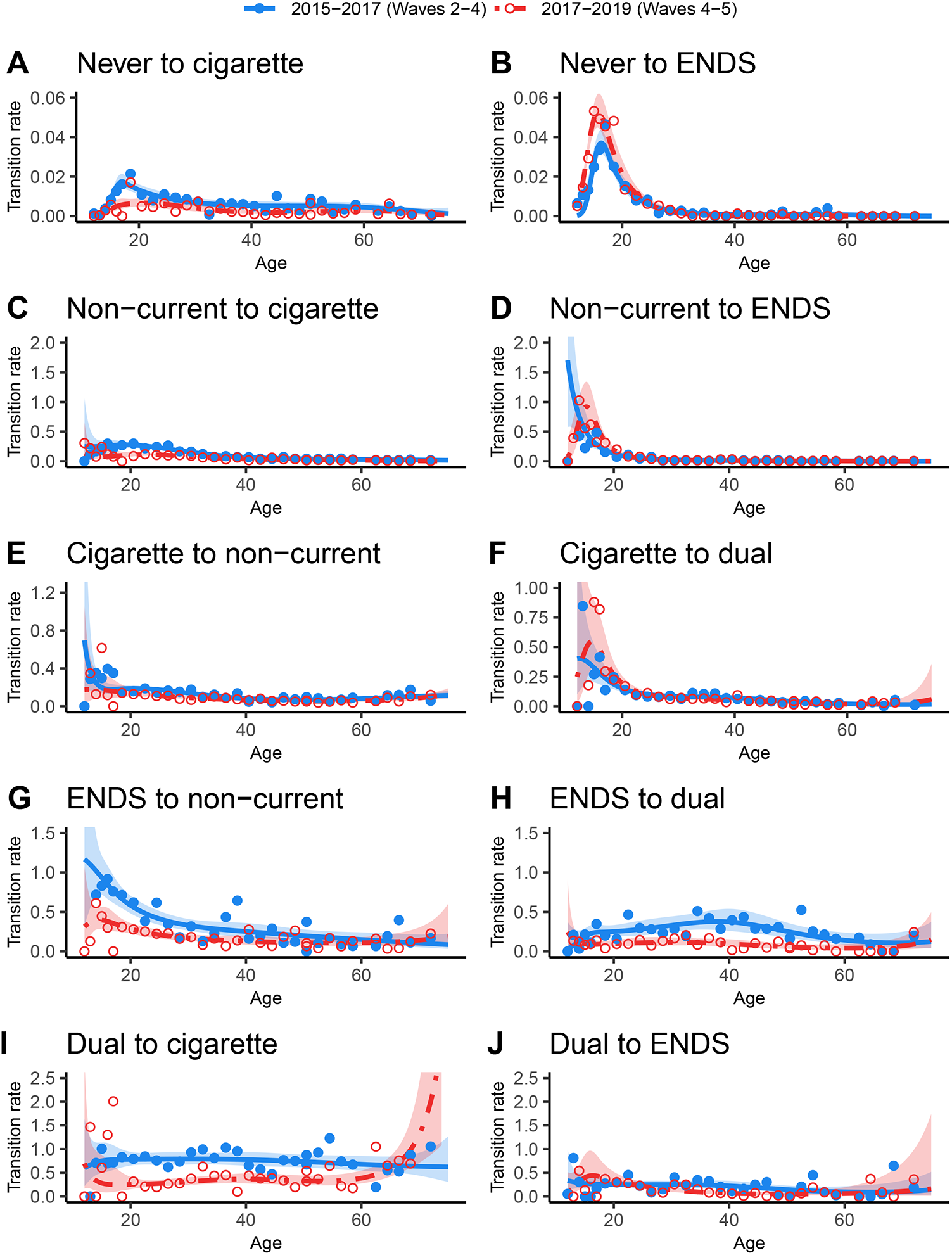
Transition rates as a function of age. Points are observed rates for participants in small age ranges, and the lines are continuous splines. Because overall transition rates depend on the persistence of each product use state, the vertical scale of each graph depends on the starting use state.

## Discussion

We estimated how patterns of initiation, cessation, and tobacco product switching changed between 2015–2017 and 2017–2019 for different age groups of youth and adults. During the latter period, there was an increase in ENDS sales driven by the uptake of JUUL and other pod-based nicotine salt systems [2]. ENDS use became more persistent, both for ENDS-only and dual cigarette and ENDS users, consistent with findings from Kasza et al. [20]. However, among adults, there has not been an increase in transitions from cigarette-only to ENDS-only or dual use. Moreover, adult dual users are more likely to remain so (instead of transitioning to cigarette-only use) and overall have not become more likely to transition to ENDS-only use. However, there are important heterogeneities in ENDS use across the adulthood spectrum [26], from young adult to middle-aged adult to older adult, with young adult ever cigarette users increasingly initiating ENDS. At the same time, youth of all user categories became more likely to transition to ENDS-only use, including dual users.

Evidence from clinical trials has suggested the ENDS can be used as a smoking cessation aid, which could have a potentially large positive public health impact [5–8]. However, evidence has been mixed about whether ENDS facilitate smoking cessation when used outside of a clinical trial context [9– 11]. We found little evidence of large changes in the probability of adult cigarette-only users initiating ENDS or transitioning to ENDS-only use (regardless of age group). Our previous analysis suggested that dual use is a typical transitional step between cigarette-only and ENDS-only use for those who do ultimately transition to full substitution of ENDS for cigarettes [15]. Our results here show that adult dual users are increasingly remaining dual users and that middle-aged and older adults have not become more likely to transition to ENDS-only use. Previous analyses of PATH Study data have raised concerns about smoking relapse among ENDS users [27, 28], which may suggest that continued nicotine addiction through ENDS use promotes dual use. However, our analysis also shows a decrease in the probability that youth or adult ENDS-only users transition to either dual or cigarette-only use. So, while most cigarette-only users are not transitioning to dual or ENDS-only use and dual users are not transitioning to ENDS-only use, ENDS-only users are increasingly less likely to transition to dual or cigarette-only use. ENDS-only users are becoming more settled in their use over time. There is substantial heterogeneity among dual users [22], so future research should seek to better understand how the ENDS initiation among cigarette users impacts frequency and intensity of smoking, as well as cessation.

There were large differences by age group among adults for ENDS-related product use transitions, with transitions more likely among younger adults. While middle and older adults have not become more likely to transition from dual use to ENDS-only use, younger adults have become more likely to completely substitute ENDS for cigarettes. We also found little evidence for increased ENDS initiation over time among adults, with the exception of young adults; ENDS use among middle-age and older adults appears to be almost entirely among ever cigarette users. We also found that young adults relapsing from non-current use are increasingly likely to use ENDS rather than cigarettes. Our results emphasize that pooling all adults together based on an age-18 cutoff masks critical, public-health-relevant heterogeneities in behavior across the adult developmental spectrum.

Unlike among adults overall, there was an increase in ENDS initiation among youth never users, roughly doubling the fraction of never users initiating regular ENDS use in one year (to nearly 4% overall). Though not high and decreasing over the time periods we considered, we did find some probability of youth transitioning from ENDS-only use to cigarette use (only or dual); ENDS use became more persistent, but the likelihood of transition to dual use did not increase. This result is consistent with other work that has found no increase in youth cigarette use [12], although lack of an increase does not mean that ENDS are not slowing the decreasing trend in cigarette use.

Our results have important implications for both researchers and regulators. Empirical studies of transitions at one time period may not be relevant at another time period, and studies, especially those that incorporate multiple years of data, should consider whether their underlying transition rates may be time-varying. For example, research developing model-based projections calibrated to transition rates estimated for specific years may not provide good projections because of this time variation, and so potential time variation should be recognized as a limitation in modeling studies. Similarly, policy recommendations can be based on both historical and the most recently available data, accounting for previous changes over time and recognize uncertainty in how transitions may continue to change in the future.

The strengths of this analysis are in the high-quality, nationally representative data of the PATH study on both youth and adults and in the multistate modeling framework that allows observed transitions to be connected to underlying transition rates. However, this work represents only a snapshot of transition behaviors in two time periods, 2015–2017 and 2017–2019. It is unclear how transition behaviors will change after this period, given the EVALI outbreak of late 2019 [29], the COVID-19 pandemic [30], the shifts in the tobacco and nicotine product marketplace [1, 2, 31], the emerging restrictions on flavored tobacco and ENDS products at the federal, state, and local level [32, 33], and the FDA decisions on millions of e-cigarette product market authorization applications (PMTA). Moreover, because PATH is a longitudinal cohort study, some of the changes in the transition rates may in part be due to individuals settling into longer term patterns. This concern should be somewhat alleviated by the replenishment in Wave 4 and the aging of shadow youth into the cohort. Other limitations of the method include the lack of accounting for individuals’ longer-term product trajectories (Markov assumption) and our inability to make causal inferences (e.g., differences in smoking initiation between never and ENDS-only users may be caused by underlying demographic differences in the two populations, not the ENDS use itself). We also did not account for ENDS flavoring in this analysis.

Our work highlights both the difference in ENDS use patterns by age, with uptake primarily among youth and young adults, and in changes between 2015–17 and 2017–2019, which may be attributed to changes in the ENDS marketplace and the ability of products to deliver nicotine [13, 34]. Newer generations of ENDS products have higher nicotine content without being unpalatable and are more efficient at nicotine delivery, which likely explains the increased persistence of ENDS use since 2017. However, adults who use ENDS, particularly older adults (ages 55+), appear to be at a higher likelihood of remaining dual users and not quitting cigarettes. Younger adult (ages 18-24) dual users have been more successful over time at transitioning to ENDS-only use, and non-current young adult users are increasingly relapsing to ENDS rather than cigarette use. Youth never users are initiating ENDS use more in 2017–2019 than in 2015–2017, but this does not appear to have increased smoking initiation. Altogether, these results reveal a complicated picture of the potential public health impact of ENDS in the United States.

While the long-term public health impact of ENDS remains to be seen, it will be shaped by public perceptions of ENDS, changes in the tobacco and nicotine product marketplace and marketing strategies, and changes in the regulatory environment. Major national public education campaigns led by the FDA and public health advocacy organizations have highlighted the dangers of ENDS use, while the ENDS industry has marketed their products’ purported benefits, influencing both youth and adult harm perceptions of ENDS. The prevention of ENDS initiation among never cigarette users is an important regulatory goal, but so is the promotion of harm reduction among current cigarette users who are not yet willing or able to quit smoking completely. US Food and Drug Administration has recently deemed some tobacco-flavored ENDS “appropriate for the protection of public health”, while issuing market denial orders for most flavored ENDS products. These regulatory actions will impact the availability of ENDS in the US marketplace and ultimately impact transitions in tobacco and nicotine product use in the future.

## What this paper adds

- Previous analyses showed that electronic nicotine delivery systems (ENDS) use was less persistent than cigarette use before 2018, but more recent studies have suggested that ENDS use is becoming more persistent.
- Observed probabilities of transitions between cigarettes and ENDS use patterns are interdependent, so underlying transition hazard rates need to be estimated to infer changes in transition propensities.
- We estimated how transition rates and probabilities changed from 2015-2017 to 2017-2019. ENDS and dual use of ENDS and cigarettes became more persistent for both adults and youth.
- Middle-aged and older adult dual users have become less likely to transition to cigarette-only use but not more likely to discontinue cigarettes, but youth and young adults have become more likely to transition to ENDS-only use.

## Data Availability

Public Use Files from the PATH study are available for download from an open access repository (https://doi.org/10.3886/ICPSR36498.v17). Restricted-Use Files (https://doi.org/ 10.3886/ICPSR36231.v31) require a Restricted Data Use Agreement. Conditions of use are available on the websites above.

https://doi.org/10.3886/ICPSR36231.v31

## Acknowledgments

This project was funded through National Cancer Institute (NCI) and Food and Drug Administration (FDA) grant U54CA229974. The opinions expressed in this article are the authors’ own and do not reflect the views of the National Institutes of Health, the Department of Health and Human Services, or the United States government.

## Data availability statement

Public Use Files from the PATH study are available for download from an open access repository (https://doi.org/10.3886/ICPSR36498.v17). Restricted-Use Files (https://doi.org/10.3886/ICPSR36231.v31) require a Restricted Data Use Agreement. Conditions of use are available on the websites above.

## Supplementary material

- Table S1: Characteristics of adults and youth in the Population Assessment of Tobacco and Health (PATH) study in 2015–2017 (Waves 2–4) and 2017–2019 (Waves 4–5), given as numbers (N) and weighted percentages (%).
- Figure S1. A) Tobacco use state definitions. (B) The direct transitions allowed between states in the model.
- Figure S2. One-year transition probabilities and confidence intervals for adults and youth in 2015–2017 (Waves 2–4) and 2017–2019 (Waves 4–5).
- Figure S3: One-year transition probabilities and confidence intervals by age group in 2015–2017 (Waves 2–4) and 2017–2019 (Waves 4–5)

**Table 1:** Covariate hazard ratios for adults and youth in 2015–2017 and 2017–2019 in multivariable multistate transition models.

|  | Male vs female | Age 12-14 vs 15-17 years | Age 18-24 vs 35-54 years | Age 25-34 vs 35-54 years | Age 55-90 vs 35-54 years | Non-Hispanic Black vs Non-Hispanic White | Hispanic vs Non-Hispanic White | Non-daily vs daily cigarette use | Non-Daily vs daily ENDS use |
| --- | --- | --- | --- | --- | --- | --- | --- | --- | --- |
| Never to cigarette |  |  |  |  |  |  |  |  |  |
| Adults 2015–2017 | **1.74 (1.29, 2.35)** | — | **2.13 (1.41, 3.23)** | 1.32 (0.84, 2.06) | 0.71 (0.43, 1.16) | **2.97 (1.92, 4.61)** | **3.61 (2.31, 5.65)** | — | — |
| Adults 2017–2019 | 1.23 (0.69, 2.20) | — | **2.58 (1.32, 5.05)** | 2.11 (0.84, 5.31) | 0.81 (0.25, 1.87) | 0.69 (0.32, 1.50) | 1.93 (0.97, 3.86) | — | — |
| Youth 2015–2017 | 0.63 (0.24, 1.62) | **0.26 (0.08, 0.91)** | — | — | — | 0.98 (0.39, 2.48) | + | — | — |
| Youth 2017–2019 | 0.42 (0.12, 1.56) | **0.26 (0.07, 0.99)** | — | — | — | 0.59 (0.20, 1.78) | + | — | — |
| Never to ENDS |  |  |  |  |  |  |  |  |  |
| Adults 2015–2017 | **2.61 (1.63, 4.19)** | — | **20.1 (4.59, 87.6)** | 4.18 (1.02, 17.1) | + | 1.31 (0.74, 2.30) | 1.12 (0.53, 2.37) | — | — |
| Adults 2017–2019 | **1.97 (1.41, 2.76)** | — | **82.2 (13.3, 509)** | 8.95 (1.31, 61.3) | + | **0.57 (0.37, 0.88)** | 0.74 (0.35, 1.59) | — | — |
| Youth 2015–2017 | **1.83 (1.19, 2.81)** | 0.30 (0.05, 1.78) | — | — | — | **0.55 (0.34, 0.91)** | + | — | — |
| Youth 2017–2019 | **1.44 (1.17, 1.78)** | **0.26 (0.20, 0.33)** | — | — | — | **0.35 (0.26, 0.46)** | **0.33 (0.19, 0.59)** | — | — |
| Non-current to cigarette |  |  |  |  |  |  |  |  |  |
| Adults 2015–2017 | 0.90 (0.73, 1.10) | — | **4.77 (3.66, 6.21)** | **3.02 (2.38, 3.85)** | **0.40 (0.29, 0.56)** | 1.08 (0.85, 1.37) | 1.19 (0.91, 1.55) | — | — |
| Adults 2017–2019 | 0.85 (0.68, 1.07) | — | **3.20 (2.40, 4.27)** | **2.60 (1.94, 3.49)** | **0.34 (0.24, 0.47)** | 1.16 (0.76, 1.76) | **1.58 (1.14, 2.20)** | — | — |
| Youth 2015–2017 | + | + | — | — | — | + | + | — | — |
| Youth 2017–2019 | 1.12 (0.48, 2.60) | 2.35 (0.61, 9.14) | — | — | — | 0.87 (0.22, 3.53) | + | — | — |
| Non-current to ENDS |  |  |  |  |  |  |  |  |  |
| Adults 2015–2017 | 0.98 (0.60, 1.62) | — | **9.48 (5.48, 16.4)** | **2.24 (1.13, 4.47)** | **0.25 (0.11, 0.54)** | 1.03 (0.55, 1.94) | **2.16 (1.00, 4.66)** | — | — |
| Adults 2017–2019 | 0.81 (0.52, 1.25) | — | **22.2 (14.0, 35.2)** | **2.98 (1.80, 4.95)** | 0.36 (0.12, 1.10) | 0.88 (0.50, 1.53) | 0.90 (0.42, 1.94) | — | — |
| Youth 2015–2017 | 1.02 (0.29, 3.57) | + | — | — | — | + | + | — | — |
| Youth 2017–2019 | 2.28 (0.90, 5.75) | 0.43 (0.09, 2.11) | — | — | — | 0.58 (0.26, 1.32) | + | — | — |
| Cigarette to non-current |  |  |  |  |  |  |  |  |  |
| Adults 2015–2017 | 1.01 (0.85, 1.21) | — | **1.75 (1.44, 2.13)** | **1.49 (1.23, 1.82)** | 1.22 (0.98, 1.54) | 1.07 (0.85, 1.35) | **0.67 (0.53, 0.85)** | **4.69 (4.08, 5.39)** | — |
| Adults 2017–2019 | 1.01 (0.84, 1.22) | — | **1.49 (1.14, 1.96)** | 1.16 (0.94, 1.45) | 1.11 (0.85, 1.45) | 1.06 (0.83, 1.36) | **0.57 (0.43, 0.76)** | **4.32 (3.66, 5.11)** | — |
| Youth 2015–2017 | 1.36 (0.24, 7.67) | + | — | — | — | + | 0.92 (0.17, 4.88) | 2.02 (0.42, 9.66) | — |
| Youth 2017–2019 | 0.73 (0.11, 4.75) | + | — | — | — | 1.12 (0.12, 10.3) | + | + | — |
| Cigarette to dual |  |  |  |  |  |  |  |  |  |
| Adults 2015–2017 | 0.90 (0.72, 1.14) | — | **2.50 (1.65, 3.76)** | **1.64 (1.25, 2.15)** | **0.40 (0.26, 0.61)** | **0.68 (0.47, 0.98)** | **0.48 (0.32, 0.72)** | + | — |
| Adults 2017–2019 | 0.90 (0.71, 1.15) | — | **3.17 (2.20, 4.55)** | **1.57 (1.02, 2.41)** | **0.38 (0.21, 0.70)** | 0.75 (0.47, 1.20) | 0.61 (0.33, 1.14) | + | — |
| Youth 2015–2017 | + | + | — | — | — | + | + | + | — |
| Youth 2017–2019 | + | + | — | — | — | + | + | + | — |
| ENDS to non-current |  |  |  |  |  |  |  |  |  |
| Adults 2015–2017 | **0.60 (0.43, 0.83)** | — | **2.16 (1.46, 3.20)** | 1.05 (0.68, 1.59) | **0.42 (0.23, 0.77)** | **2.32 (1.69, 3.18)** | **2.67 (1.63, 4.39)** | — | 1.13 (0.22, 5.79) |
| Adults 2017–2019 | 0.80 (0.56, 1.13) | — | **2.20 (1.33, 3.64)** | 1.32 (0.73, 2.39) | 1.01 (0.52, 1.98) | 1.43 (0.89, 2.28) | 1.59 (0.89, 2.85) | — | 1.39 (0.61, 3.18) |
| Youth 2015–2017 | 1.14 (0.46, 2.79) | 1.42 (0.22, 9.2) | — | — | — | + | + | — | **9.52 (1.56, 58.0)** |
| Youth 2017–2019 | 1.66 (0.94, 2.92) | **1.84 (1.07, 3.17)** | — | — | — | 1.00 (0.48, 1.71) | 1.86 (0.28, 12.5) | — | + |
| ENDS to dual |  |  |  |  |  |  |  |  |  |
| Adults 2015–2017 | 1.25 (0.87, 1.83) | — | 0.82 (0.48, 1.39) | 0.78 (0.48, 1.24) | **0.41 (0.21, 0.83)** | 0.85 (0.49, 1.49) | 0.61 (0.24, 1.52) | — | + |
| Adults 2017–2019 | 0.97 (0.60, 1.55) | — | 1.20 (0.62, 2.34) | 1.54 (0.69, 3.42) | 0.67 (0.24, 1.87) | 0.99 (0.52, 1.89) | + | — | + |
| Youth 2015–2017 | 0.90 (0.24, 3.28) | + | — | — | — | 0.72 (0.14, 3.84) | + | — | + |
| Youth 2017–2019 | 1.49 (0.58, 3.87) | + | — | — | — | + | 3.47 (0.64, 18.6) | — | + |
| Dual to cigarette |  |  |  |  |  |  |  |  |  |
| Adults 2015–2017 | 1.07 (0.87, 1.32) | — | 1.18 (0.90, 1.57) | 0.98 (0.76, 1.26) | 0.84 (0.64, 1.09) | 1.17 (0.83, 1.64) | 0.98 (0.62, 1.55) | **0.57 (0.41, 0.80)** | 0.98 (0.69, 1.41) |
| Adults 2017–2019 | 0.90 (0.63, 1.26) | — | 0.73 (0.47, 1.14) | 0.93 (0.60, 1.46) | 1.42 (0.92, 2.19) | 1.11 (0.71, 1.74) | 0.99 (0.55, 1.77) | 0.97 (0.59, 1.58) | **0.53 (0.36, 0.78)** |
| Youth 2015–2017 | + | + | — | — | — | + | + | + | + |
| Youth 2017–2019 | + | + | — | — | — | + | + | + | + |
| Dual to ENDS |  |  |  |  |  |  |  |  |  |
| Adults 2015–2017 | 0.98 (0.49, 1.95) | — | 1.32 (0.64, 2.71) | 0.94 (0.50, 1.76) | 0.57 (0.16, 2.07) | 0.91 (0.48, 1.71) | 0.87 (0.27, 2.82) | + | + |
| Adults 2017–2019 | 1.01 (0.55, 1.87) | — | **5.48 (2.40, 12.5)** | **3.37 (1.28, 8.89)** | 1.62 (0.41, 6.43) | 0.65 (0.24, 1.76) | 1.47 (0.49, 4.40) | 1.20 (0.43, 3.33) | **3.70 (1.29, 10.6)** |
| Youth 2015–2017 | + | + | — | — | — | + | + | + | + |
| Youth 2017–2019 | + | + | — | — | — | + | + | + | + |

†Estimate suppressed because of high variance (variance of log(HR)>1)

## References

[1] Huang J, Duan Z, Kwok J, Binns S, Vera LE, Kim Y, et al. Vaping versus JUULing: how the extraordinary growth and marketing of JUUL transformed the US retail e-cigarette market. Tobacco control. 2019;28(2):146–51.

[2] Ali FRM, Diaz MC, Vallone D, Tynan MA, Cordova J, Seaman EL, et al. E-cigarette unit sales, by product and flavor type—United States, 2014–2020. Morbidity and Mortality Weekly Report. 2020;69(37):1313.

[3] Hammond D, Reid JL, Burkhalter R, O’Connor RJ, Goniewicz ML, Wackowski OA, et al. Trends in e-cigarette brands, devices and the nicotine profile of products used by youth in England, Canada and the USA: 2017–2019. Tobacco control. 2021.

[4] Farzal Z, Perry MF, Yarbrough WG, Kimple AJ. The adolescent vaping epidemic in the United States—How it happened and where we go from here. JAMA Otolaryngology–Head & Neck Surgery. 2019;145(10):885–6.

[5] Bullen C, Howe C, Laugesen M, McRobbie H, Parag V, Williman J, et al. Electronic cigarettes for smoking cessation: a randomised controlled trial. The Lancet. 2013;382(9905):1629–37.

[6] Hajek P. Electronic cigarettes have a potential for huge public health benefit. BMC Medicine. 2014;12(1):1–4.

[7] McNeill A. Should clinicians recommend e-cigarettes to their patients who smoke? Yes. Annals of Family Medicine. 2016;14(4):300–1.

[8] Hartmann-Boyce J, McRobbie H, Butler AR, Lindson N, Bullen C, Begh R, et al. Electronic cigarettes for smoking cessation. Cochrane database of systematic reviews. 2021;(9).

[9] Kalkhoran S, Glantz SA. E-cigarettes and smoking cessation in real-world and clinical settings: A systematic review and meta-analysis. The Lancet Respiratory Medicine. 2016;4(2):116–28.

[10] Bhatnagar A, Payne TJ, Robertson RM. Is There A Role for Electronic Cigarettes in Tobacco Cessation? Journal of the American Heart Association. 2019;8(12):10–3.

[11] Wang RJ, Bhadriraju S, Glantz SA. E-Cigarette Use and Adult Cigarette Smoking Cessation: A Meta-Analysis. American Journal of Public Health. 2021;111(2):230–46.

[12] Johnston, L. D., Miech, R. A., O’Malley, P. M., Bachman, J. G., Schulenberg, J. E., & Patrick, M. E. (2022). Monitoring the Future national survey results on drug use 1975-2021: Overview, key findings on adolescent drug use. Ann Arbor: Institute for Social Research, University of Michigan.

[13] Leventhal AM, Madden DR, Peraza N, Schiff SJ, Lebovitz L, Whitted L, et al. Effect of exposure to e-cigarettes with salt vs free-base nicotine on the appeal and sensory experience of vaping: A randomized clinical trial. JAMA Network Open. 2021;4(1):e2032757–7.

[14] McKelvey K, Baiocchi M, Ramamurthi D, McLaughlin S, Halpern-Felsher B. Youth say ads for flavored e-liquids are for them. Addictive behaviors.2019;91:164–70.

[15] Brouwer AF, Jeon J, Hirschtick JL, Jimenez-Mendoza E, Mistry R, Bondarenko IV, et al. Transitions between cigarette, ENDS and dual use in adults in the PATH study (waves 1–4): multistate transition modelling accounting for complex survey design. Tobacco Control. 2022;31(3):424–31.

[16] Brouwer AF, Jeon J, Cook SF, Usidame B, Hirschtick JL, Jimenez-Mendoza E, et al. The Impact of Menthol Cigarette Flavor in the U.S.: Cigarette and ENDS Transitions by Sociodemographic Group. American Journal of Preventive Medicine. 2022;62(2):243–51.

[17] Shafie-Khorassani F, Piper ME, Jorenby DE, Baker TB, Benowitz NL, Hayes-Birchler T, Meza R, Brouwer AF. Associations of demographics, dependence, and biomarkers with transitions in tobacco product use in a cohort of cigarette users and dual users of cigarettes and e-cigarettes. Nicotine & Tobacco Research. 2022.

[18] Mantey DS, Harrell MB, Chen B, Kelder SH, Perry CL, Loukas A. A Longitudinal Examination of Behavioral Transitions among Young Adult Menthol and Non-Menthol Cigarette Smokers Using a Three-State Markov Model. Nicotine and Tobacco Research. 2021;23(6):1047–54.

[19] Loukas A, Marti CN, Harrell MB. Electronic nicotine delivery systems use predicts transitions in cigarette smoking among young adults. Drug and Alcohol Dependence. 2022;231:109251.

[20] Kasza KA, Tang Z, Xiao H, Marshall D, Stanton C, Gross AL, et al. National longitudinal tobacco product cessation rates among US adults from the PATH Study: 2013–2019 (waves 1–5). Tobacco Control. 2022.

[21] National Institute on Drug Abuse, Food and Drug Administration Center for Tobacco Products. Population Assessment of Tobacco and Health (PATH) Study [United States] Public-Use Files; 2022. Inter-university Consortium for Political and Social Research [distributor], 2019-11-21.

[22] National Institute on Drug Abuse, Food and Drug Administration Center for Tobacco Products. Population Assessment of Tobacco and Health (PATH) Study [United States] Restricted-Use Files; 2022. Inter-university Consortium for Political and Social Research [distributor], 2022-11-19.

[23] Brouwer AF, Levy DT, Jeon J, Jimenez-Mendoza E, Sanchez-Romero LM, Mistry R, et al. The impact of current tobacco product use definitions on estimates of transitions between cigarette and ENDS use. Nicotine & Tobacco Research. 2022.

[24] Durrett R. Essentials of Stochastic Processes. Springer; 1999.

[25] Wang W, Yan J. Shape-Restricted Regression Splines with R Package splines2. Journal of Data Science. 2021; 19(3):498–517.

[26] Alexander CN, Langer EJ. Higher stages of human development: Perspectives on adult growth. Oxford University Press; 1990.

[27] Azagba S, Qeadan F, Shan L, Latham K, Wolfson M. E-Cigarette Use and Transition in Adult Smoking Frequency: A Longitudinal Study. American Journal of Preventive Medicine. 2020; 59(3):367–76.

[28] Everard CD, Silveira ML, Kimmel HL, Marshall D, Blanco C, Compton WM. Association of Electronic Nicotine Delivery System Use With Cigarette Smoking Relapse Among Former Smokers in the United States. JAMA Network Open. 2020;3(6):e204813.

[29] Krishnasamy VP, Hallowell BD, Ko JY, Board A, Hartnett KP, Salvatore PP, et al. Update: characteristics of a nationwide outbreak of e-cigarette, or vaping, product use–associated lung injury—United States, August 2019– January 2020. Morbidity and Mortality Weekly Report. 2020;69(3):90.

[30] White AM, Li D, Snell LM, O’Connor R, Hoetger C, Croft D, et al. Perceptions of tobacco product-specific COVID-19 risk and changes in tobacco use behaviors among smokers, e-cigarette users, and dual users. Nicotine and Tobacco Research. 2021;23(9):1617–22.

[31] Hammond D, Reid JL, Burkhalter R, Bansal Travers M, Gravely S, Hyland A, et al. E-Cigarette Flavors, Devices, and Brands Used by Youths Before and After Partial Flavor Restrictions in the United States: Canada, England, and the United States, 2017–2020. American Journal of Public Health. 2022;112(7):1014–24.

[32] Rose SW, Amato MS, Anesetti-Rothermel A, Carnegie B, Safi Z, Benson AF, Czaplicki L, Simpson R, Zhou Y, Akbar M, Younger Gagosian S. Characteristics and reach equity of policies restricting flavored tobacco product sales in the United States. Health Promotion Practice. 2020;21(1_suppl):44S–53S.

[33] Gravely S, Smith DM, Liber AC, Cummings KM, East KA, Hammond D, Hyland A, O’Connor RJ, Kasza KA, Quah AC, Loewen R. Responses to potential nicotine vaping product flavor restrictions among regular vapers using non-tobacco flavors: Findings from the 2020 ITC Smoking and Vaping Survey in Canada, England and the United States. Addictive Behaviors. 2022;125:107152.

[34] Gholap VV, Kosmider L, Golshahi L, Halquist MS. Nicotine forms: why and how do they matter in nicotine delivery from electronic cigarettes?. Expert opinion on drug delivery. 2020;17(12):1727–36.

