## Supplementary material for "Changing patterns of cigarette and ENDS transitions in the US: a multistate transition analysis of youth and adults in the PATH Study in 2015–17 vs 2017–2019": Table S1

|  | Youth Waves 2-4 | | Youth Waves 4-5 | | Adults Waves 2-4 | | Adults Waves 4-5 | |
| --- | --- | --- | --- | --- | --- | --- | --- | --- |
|  | % | N | % | N | % | N | % | N |
| Total | 100 | 12067 | 100 | 12538 | 100 | 24242 | 100 | 28061 |
| Gender |  |  |  |  |  |  |  |  |
| Female | 48.6 | 5861 | 49.0 | 6054 | 52.1 | 12591 | 62.0 | 13522 |
| Male | 51.3 | 6206 | 51.0 | 6484 | 47.9 | 11651 | 48.0 | 14539 |
| Race/ethnicity |  |  |  |  |  |  |  |  |
| Non-Hispanic White | 51.9 | 5509 | 51.2 | 5528 | 64.3 | 13991 | 63.7 | 15840 |
| Non-Hispanic Black | 22.9 | 3504 | 22.9 | 3741 | 15.3 | 4544 | 15.5 | 5505 |
| Hispanic | 12.7 | 1586 | 12.7 | 1635 | 11.1 | 3543 | 11.0 | 4171 |
| Non-Hispanic Other/Unknown | 12.5 | 1468 | 13.2 | 1634 | 9.3 | 2164 | 9.8 | 2545 |
| Age (years) |  |  |  |  |  |  |  |  |
| 12-14 | 57.5 | 7123 | 49.2 | 6254 |  |  |  |  |
| 15-17 | 42.5 | 4944 | 50.8 | 6284 |  |  |  |  |
| 18-24 |  |  |  |  | 14.3 | 7929 | 12.2 | 8891 |
| 25-34 |  |  |  |  | 17.5 | 4581 | 17.9 | 5794 |
| 35-54 |  |  |  |  | 33.2 | 6719 | 33.3 | 7470 |
| 55+ |  |  |  |  | 35.0 | 5013 | 36.5 | 5906 |
| Tobacco & ENDS use state |  |  |  |  |  |  |  |  |
| Never user | 96.6 | 11692 | 96.2 | 12066 | 58.8 | 11968 | 57.9 | 13894 |
| Non-current user | 1.1 | 127 | 1.4 | 183 | 21.8 | 4079 | 23.1 | 5323 |
| Cigarette-only user | 1.0 | 116 | 0.7 | 92 | 16.4 | 6875 | 16.0 | 7365 |
| Non-daily | 0.5 | 59 | 0.4 | 49 | 3.4 | 1320 | 3.2 | 1425 |
| Daily | 0.5 | 57 | 0.3 | 43 | 13.1 | 5555 | 12.8 | 5940 |
| ENDS-only user | 1.0 | 99 | 1.4 | 169 | 1.3 | 564 | 1.5 | 758 |
| Non-daily | 0.8 | 77 | 1.2 | 144 | 0.4 | 208 | 0.4 | 249 |
| Daily | 0.2 | 22 | 0.2 | 25 | 0.9 | 356 | 1.1 | 509 |
| Dual cigarette/ENDS user | 0.3 | 33 | 0.2 | 28 | 1.7 | 756 | 1.5 | 721 |
| Non-daily cigarette, non-daily ENDS | 0.1 | 16 | 0.1 | 16 | 0.2 | 92 | 0.2 | 100 |
| Non-daily cigarette, daily ENDS | 0.1 | 6 | 0.0 | 3 | 0.3 | 142 | 0.4 | 168 |
| Daily cigarette, non-daily ENDS | 0.1 | 8 | 0.0 | 5 | 0.8 | 362 | 0.6 | 316 |
| Daily cigarette, daily ENDS | 0.0 | 3 | 0.0 | 4 | 0.4 | 160 | 0.3 | 137 |

**Table S1**: Characteristics of adults and youth in the Population Assessment of Tobacco and Health (PATH) study in 2015–2017 (Waves 2–4) and 2017–2019 (Waves 4–5), given as numbers (N) and weighted percentages (%).
