## Supplementary figures and images for "Changing patterns of cigarette and ENDS transitions in the US: a multistate transition analysis of youth and adults in the PATH Study in 2015–17 vs 2017–2019"

### Figure S1

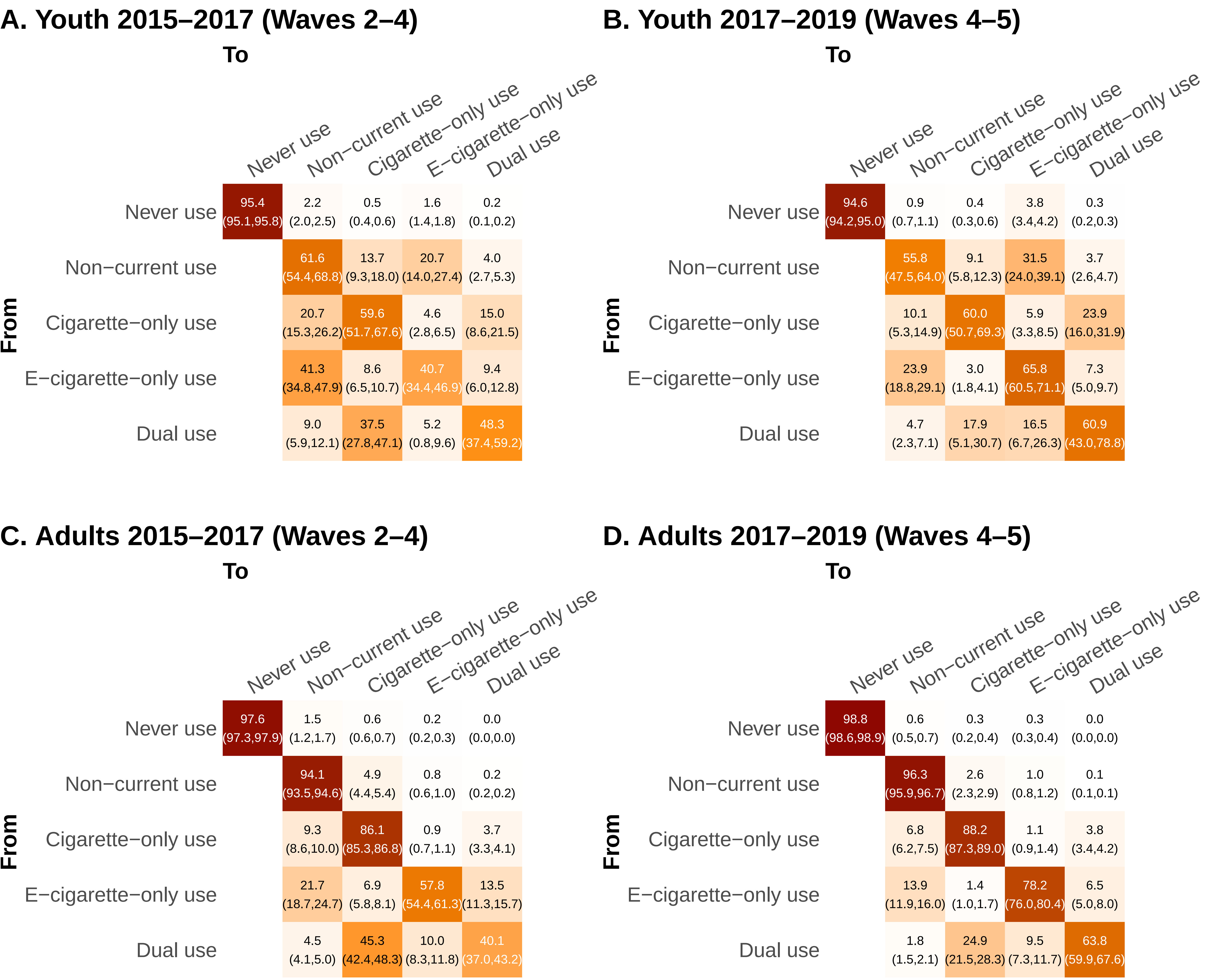

### Figure S3

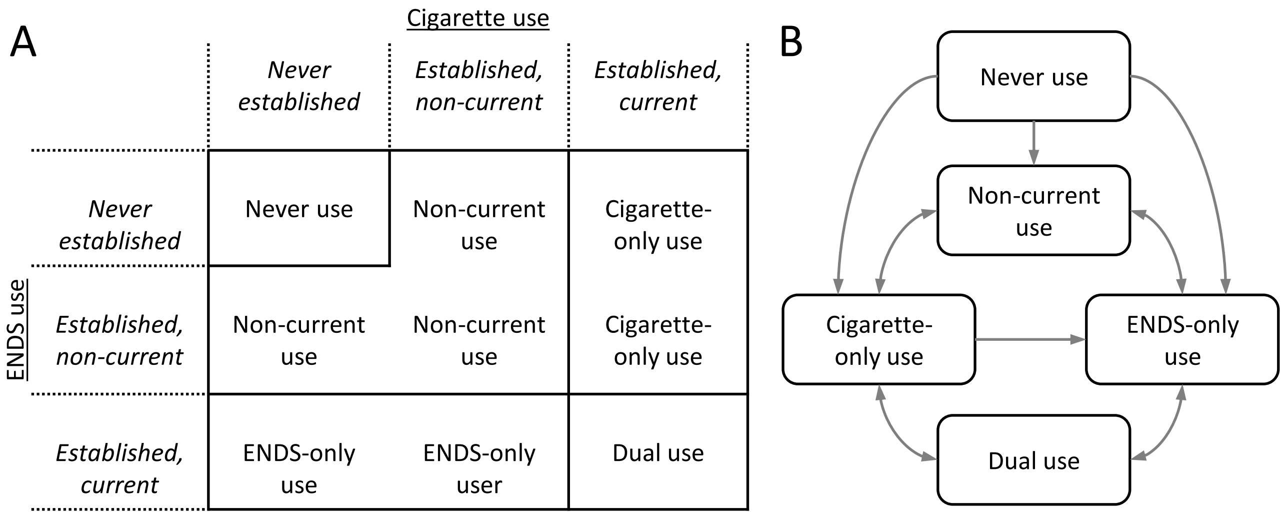
