## Supplementary material for "Changing patterns of cigarette and ENDS transitions in the US: a multistate transition analysis of youth and adults in the PATH Study in 2015–17 vs 2017–2019": Figure S2

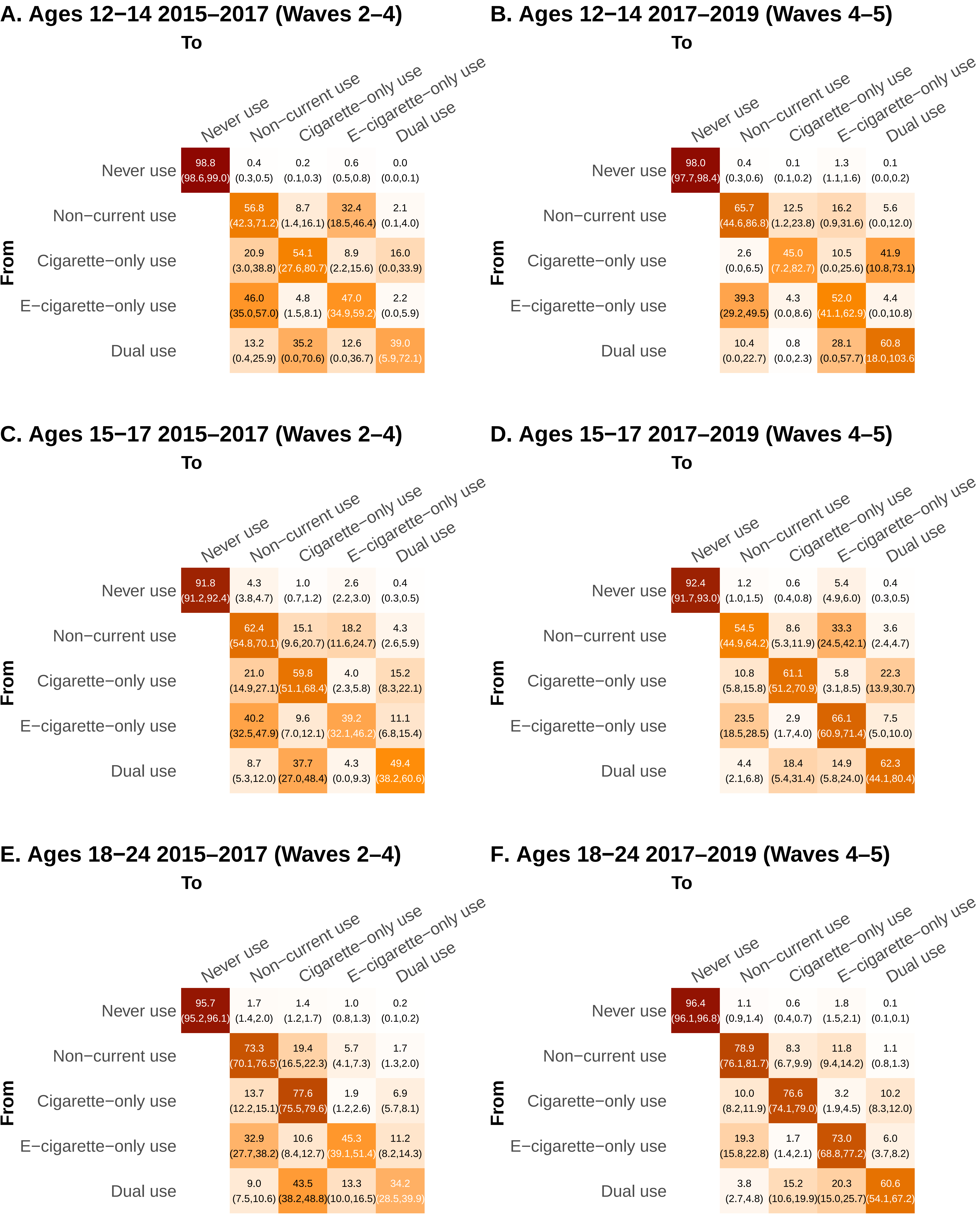
A. Ages 12−14 2015−2017 (Waves 2−4)
To
From
Never use
Non−current use
Cigarette−only use
E−cigarette−only use
Dual use
Never use: 98.8 (98.6,99.0) | 0.4 (0.3,0.5) | 0.2 (0.1,0.3) | 0.6 (0.5,0.8) | 0.0 (0.0,0.1)
Non−current use: 56.8 (42.3,71.2) | 8.7 (1.4,16.1) | 32.4 (18.5,46.4) | 2.1 (0.1,4.0)
Cigarette−only use: 20.9 (3.0,38.8) | 54.1 (27.6,80.7) | 8.9 (2.2,15.6) | 16.0 (0.0,33.9)
E−cigarette−only use: 46.0 (35.0,57.0) | 4.8 (1.5,8.1) | 47.0 (34.9,59.2) | 2.2 (0.0,5.9)
Dual use: 13.2 (0.4,25.9) | 35.2 (0.0,70.6) | 12.6 (0.0,36.7) | 39.0 (5.9,72.1)
B. Ages 12−14 2017−2019 (Waves 4−5)
To
From
Never use
Non−current use
Cigarette−only use
E−cigarette−only use
Dual use
Never use: 98.0 (97.7,98.4) | 0.4 (0.3,0.6) | 0.1 (0.1,0.2) | 1.3 (1.1,1.6) | 0.1 (0.0,0.2)
Non−current use: 65.7 (44.6,86.8) | 12.5 (1.2,23.8) | 16.2 (0.9,31.6) | 5.6 (0.0,12.0)
Cigarette−only use: 2.6 (0.0,6.5) | 45.0 (7.2,82.7) | 10.5 (0.0,25.6) | 41.9 (10.8,73.1)
E−cigarette−only use: 39.3 (29.2,49.5) | 4.3 (0.0,8.6) | 52.0 (41.1,62.9) | 4.4 (0.0,10.8)
Dual use: 10.4 (0.0,22.7) | 0.8 (0.0,2.3) | 28.1 (0.0,57.7) | 60.8 (18.0,103.6
C. Ages 15−17 2015−2017 (Waves 2−4)
To
From
Never use
Non−current use
Cigarette−only use
E−cigarette−only use
Dual use
Never use: 91.8 (91.2,92.4) | 4.3 (3.8,4.7) | 1.0 (0.7,1.2) | 2.6 (2.2,3.0) | 0.4 (0.3,0.5)
Non−current use: 62.4 (54.8,70.1) | 15.1 (9.6,20.7) | 18.2 (11.6,24.7) | 4.3 (2.6,5.9)
Cigarette−only use: 21.0 (14.9,27.1) | 59.8 (51.1,68.4) | 4.0 (2.3,5.8) | 15.2 (8.3,22.1)
E−cigarette−only use: 40.2 (32.5,47.9) | 9.6 (7.0,12.1) | 39.2 (32.1,46.2) | 11.1 (6.8,15.4)
Dual use: 8.7 (5.3,12.0) | 37.7 (27.0,48.4) | 4.3 (0.0,9.3) | 49.4 (38.2,60.6)
D. Ages 15−17 2017−2019 (Waves 4−5)
To
From
Never use
Non−current use
Cigarette−only use
E−cigarette−only use
Dual use
Never use: 92.4 (91.7,93.0) | 1.2 (1.0,1.5) | 0.6 (0.4,0.8) | 5.4 (4.9,6.0) | 0.4 (0.3,0.5)
Non−current use: 54.5 (44.9,64.2) | 8.6 (5.3,11.9) | 33.3 (24.5,42.1) | 3.6 (2.4,4.7)
Cigarette−only use: 10.8 (5.8,15.8) | 61.1 (51.2,70.9) | 5.8 (3.1,8.5) | 22.3 (13.9,30.7)
E−cigarette−only use: 23.5 (18.5,28.5) | 2.9 (1.7,4.0) | 66.1 (60.9,71.4) | 7.5 (5.0,10.0)
Dual use: 4.4 (2.1,6.8) | 18.4 (5.4,31.4) | 14.9 (5.8,24.0) | 62.3 (44.1,80.4)
E. Ages 18−24 2015−2017 (Waves 2−4)
To
From
Never use
Non−current use
Cigarette−only use
E−cigarette−only use
Dual use
Never use: 95.7 (95.2,96.1) | 1.7 (1.4,2.0) | 1.4 (1.2,1.7) | 1.0 (0.8,1.3) | 0.2 (0.1,0.2)
Non−current use: 73.3 (70.1,76.5) | 19.4 (16.5,22.3) | 5.7 (4.1,7.3) | 1.7 (1.3,2.0)
Cigarette−only use: 13.7 (12.2,15.1) | 77.6 (75.5,79.6) | 1.9 (1.2,2.6) | 6.9 (5.7,8.1)
E−cigarette−only use: 32.9 (27.7,38.2) | 10.6 (8.4,12.7) | 45.3 (39.1,51.4) | 11.2 (8.2,14.3)
Dual use: 9.0 (7.5,10.6) | 43.5 (38.2,48.8) | 13.3 (10.0,16.5) | 34.2 (28.5,39.9)
F. Ages 18−24 2017−2019 (Waves 4−5)
To
From
Never use
Non−current use
Cigarette−only use
E−cigarette−only use
Dual use
Never use: 96.4 (96.1,96.8) | 1.1 (0.9,1.4) | 0.6 (0.4,0.7) | 1.8 (1.5,2.1) | 0.1 (0.1,0.1)
Non−current use: 78.9 (76.1,81.7) | 8.3 (6.7,9.9) | 11.8 (9.4,14.2) | 1.1 (0.8,1.3)
Cigarette−only use: 10.0 (8.2,11.9) | 76.6 (74.1,79.0) | 3.2 (1.9,4.5) | 10.2 (8.3,12.0)
E−cigarette−only use: 19.3 (15.8,22.8) | 1.7 (1.4,2.1) | 73.0 (68.8,77.2) | 6.0 (3.7,8.2)
Dual use: 3.8 (2.7,4.8) | 15.2 (10.6,19.9) | 20.3 (15.0,25.7) | 60.6 (54.1,67.2)

### G. Ages 25−34 2015−2017 (Waves 2−4)

| From \ To | Never use | Non−current use | Cigarette−only use | E−cigarette−only use | Dual use |
|---|---|---|---|---|---|
| Never use | 97.8 95.5,100.1 | 1.2 (0.0,4.3) | 0.8 (0.1,1.5) | 0.2 (0.0,0.8) | 0.1 (0.0,0.2) |
| Non−current use | | 84.0 51.2,116.8 | 13.6 (0.0,40.6) | 1.7 (0.0,6.0) | 0.7 (0.0,2.2) |
| Cigarette−only use | | 11.6 (0.0,32.3) | 82.0 63.2,100.8 | 1.3 (0.0,2.7) | 5.1 (0.0,12.2) |
| E−cigarette−only use | | 19.4 (0.0,44.0) | 7.5 (0.8,14.2) | 59.7 (29.1,90.2) | 13.5 (0.0,28.7) |
| Dual use | | 4.9 (0.0,10.1) | 43.3 (35.3,51.3) | 9.6 (6.5,12.7) | 42.2 (35.9,48.5) |

### H. Ages 25−34 2017−2019 (Waves 4−5)

| From \ To | Never use | Non−current use | Cigarette−only use | E−cigarette−only use | Dual use |
|---|---|---|---|---|---|
| Never use | 98.8 97.6,100.1 | 0.5 (0.1,0.8) | 0.5 (0.0,1.0) | 0.2 (0.0,0.8) | 0.0 (0.0,0.1) |
| Non−current use | | 90.4 69.8,110.9 | 7.4 (0.0,22.3) | 1.9 (0.0,6.8) | 0.3 (0.0,1.2) |
| Cigarette−only use | | 7.8 (0.0,31.4) | 85.2 64.4,106.1 | 1.7 (0.0,5.0) | 5.2 (0.0,12.9) |
| E−cigarette−only use | | 13.7 (0.0,33.0) | 2.0 (0.0,4.5) | 75.6 49.6,101.5 | 8.7 (0.0,20.0) |
| Dual use | | 2.2 (0.0,4.8) | 23.0 (16.6,29.5) | 11.8 (7.1,16.6) | 63.0 (55.8,70.2) |

### I. Ages 35−54 2015−2017 (Waves 2−4)

| From \ To | Never use | Non−current use | Cigarette−only use | E−cigarette−only use | Dual use |
|---|---|---|---|---|---|
| Never use | 98.4 (98.0,98.7) | 1.0 (0.7,1.3) | 0.5 (0.4,0.7) | 0.1 (0.0,0.1) | 0.0 (0.0,0.0) |
| Non−current use | | 94.1 (93.1,95.2) | 4.9 (3.9,5.8) | 0.8 (0.4,1.1) | 0.2 (0.1,0.3) |
| Cigarette−only use | | 7.3 (6.4,8.2) | 88.7 (87.7,89.8) | 0.6 (0.3,0.9) | 3.4 (2.8,3.9) |
| E−cigarette−only use | | 20.6 (15.3,26.0) | 8.2 (6.0,10.4) | 55.2 (48.1,62.4) | 15.9 (11.7,20.1) |
| Dual use | | 3.9 (3.0,4.7) | 46.2 (40.8,51.5) | 9.8 (6.5,13.1) | 40.1 (34.7,45.5) |

### J. Ages 35−54 2017−2019 (Waves 4−5)

| From \ To | Never use | Non−current use | Cigarette−only use | E−cigarette−only use | Dual use |
|---|---|---|---|---|---|
| Never use | 99.3 (99.1,99.5) | 0.4 (0.2,0.6) | 0.2 (0.1,0.3) | 0.0 (0.0,0.1) | 0.0 (0.0,0.0) |
| Non−current use | | 96.4 (95.6,97.1) | 2.9 (2.3,3.5) | 0.7 (0.4,0.9) | 0.1 (0.1,0.1) |
| Cigarette−only use | | 6.0 (5.2,6.8) | 89.9 (88.8,91.0) | 0.7 (0.4,1.0) | 3.4 (2.7,4.1) |
| E−cigarette−only use | | 11.3 (7.2,15.3) | 1.2 (0.6,1.8) | 81.8 (76.9,86.7) | 5.7 (3.0,8.5) |
| Dual use | | 1.2 (0.8,1.5) | 26.5 (20.8,32.2) | 3.7 (1.4,5.9) | 68.7 (62.3,75.1) |

### K. Ages 55−90 2015−2017 (Waves 2−4)

| From \ To | Never use | Non−current use | Cigarette−only use | E−cigarette−only use | Dual use |
|---|---|---|---|---|---|
| Never use | 97.7 (97.1,98.3) | 2.0 (1.4,2.5) | 0.3 (0.2,0.4) | 0.0 (0.0,0.1) | 0.0 (0.0,0.0) |
| Non−current use | | 97.7 (97.2,98.2) | 2.0 (1.5,2.5) | 0.2 (0.1,0.4) | 0.0 (0.0,0.0) |
| Cigarette−only use | | 8.3 (6.9,9.8) | 89.6 (88.1,91.1) | 0.6 (0.3,0.9) | 1.5 (1.0,2.0) |
| E−cigarette−only use | | 10.1 (5.2,14.9) | 3.2 (1.5,5.0) | 78.3 (70.9,85.7) | 8.4 (4.2,12.6) |
| Dual use | | 2.6 (2.0,3.3) | 43.5 (36.6,50.5) | 6.3 (1.4,11.3) | 47.5 (39.3,55.7) |

### L. Ages 55−90 2017−2019 (Waves 4−5)

| From \ To | Never use | Non−current use | Cigarette−only use | E−cigarette−only use | Dual use |
|---|---|---|---|---|---|
| Never use | 99.3 (99.0,99.5) | 0.6 (0.3,0.8) | 0.2 (0.1,0.3) | 0.0 (0.0,0.0) | 0.0 (0.0,0.0) |
| Non−current use | | 98.8 (98.4,99.1) | 1.0 (0.7,1.3) | 0.2 (0.0,0.5) | 0.0 (0.0,0.0) |
| Cigarette−only use | | 6.7 (5.2,8.1) | 91.6 (90.1,93.2) | 0.4 (0.2,0.7) | 1.3 (0.7,1.8) |
| E−cigarette−only use | | 11.4 (6.0,16.8) | 1.0 (0.2,1.9) | 84.1 (77.8,90.5) | 3.4 (0.8,6.1) |
| Dual use | | 1.8 (1.2,2.5) | 37.1 (27.5,46.6) | 5.6 (1.0,10.3) | 55.5 (45.2,65.8) |
